# PTL-PRS: an R package for transfer learning of polygenic risk scores with pseudovalidation

**DOI:** 10.1101/2025.06.19.25329937

**Authors:** Bokeum Cho, Seunggeun Lee

## Abstract

**Summary:** Polygenic risk scores (PRSs) are essential tools for predicting individual phenotypic risk but often lack accuracy in non-European ancestry groups. Transfer Learning for Polygenic Risk Scores (TL-PRS) addresses this challenge by leveraging European PRSs to improve prediction in underrepresented ancestries but requires privacy-sensitive individual-level data and has low computational efficiency. Therefore, we introduce PTL-PRS (Pseudovalidated Transfer Learning for PRS), an extension of TL-PRS that incorporates pseudovalidation to eliminate the need for individual-level data and includes further software optimization. For pseudovalidation, PTL-PRS generates pseudo-summary statistics for training and validation and evaluates model performance with the pseudo-*R* metric. To improve computational efficiency, PTL-PRS software was optimized with C++, blockwise early stopping, and direct genotype retrieval. Overall, PTL-PRS enhances both prediction accuracy and software usability, helping underrepresented populations achieve more accurate genetic risk predictions.

**Availability and Implementation:** The *PTL*.*PRS* R package is publicly available on GitHub at https://github.com/bokeumcho/PTL.PRS. The summary statistics used in this paper are available in the public domain: UK Biobank (https://pheweb.org/UKB-TOPMed), PGS Catalog (https://www.pgscatalog.org) and GenOMICC (https://genomicc.org/data).

## Introduction

Polygenic risk scores (PRSs) estimate an individual’s genetic predisposition to traits or diseases on the basis of genome-wide association study (GWAS) data [1-3]. They have also gained widespread application in biomedical research, including the assessment of shared etiology between phenotypes [4, 5]. Despite their potential, PRSs are difficult to implement in clinical settings; for example, they do not perform as well in individuals of non-European ancestry [6-8].

To address these challenges, Transfer Learning for Polygenic Risk Scores (TL-PRS) [9] was developed. TL-PRS leverages large-scale datasets from European populations to enhance the predictive performance of PRSs in underrepresented groups, thereby improving model generalizability and reducing bias. However, its practical application is limited by the need for individual-level data [10], which are required for hyperparameter tuning and performance evaluation but are often unavailable due to privacy concerns.

To address this limitation, we introduce a pseudovalidation strategy for TL-PRS that enables model validation using only publicly available GWAS summary statistics, eliminating the need for individual-level data. This method builds upon the pseudosplitting framework established in PUMAS [10] and extended in MegaPRS [11], which partitions summary statistics into pseudotraining and pseudovalidation sets. Model performance is evaluated using the pseudo-*R* metric [11], which facilitates the effective construction and assessment of TL-PRS models under limited data availability.

We implemented this strategy in PTL-PRS (Pseudovalidated Transfer Learning for Polygenic Risk Scores), a scalable software package optimized for performance. PTL-PRS reimplements core training routines in C++ to support multithreaded parallelism and incorporates techniques such as blockwise early stopping and index-based SNP retrieval from binary genotype files. These optimizations result in substantially reduced memory consumption and faster training times relative to those of the original TL-PRS framework.

## Methods

PTL-PRS consists of three main steps with three inputs, as shown in **Figure 1**. The required inputs (part (A) in **Figure 1**) are all summary statistics, including the following:

1. Target-ancestry GWAS summary statistics (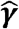: marginal SNP effect sizes, 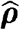: SNP□phenotype correlations),
2. Target population reference LD estimates (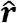: pairwise SNP correlations), and
3. Source-ancestry PRS weights (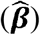).

**Figure 1.**
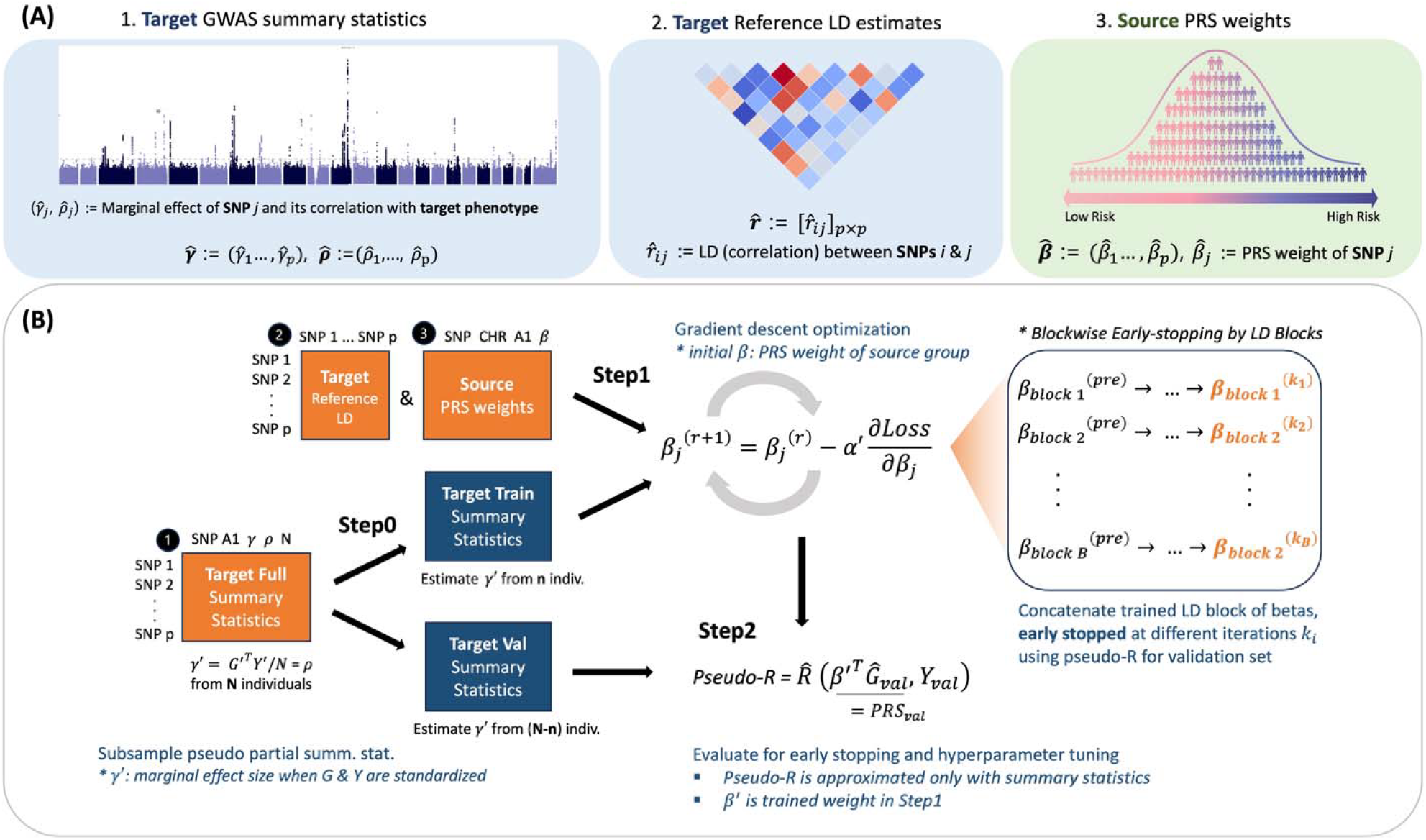
Overview of the PTL-PRS framework and its required inputs (A) **Three necessary inputs**: Target-ancestry GWAS summary statistics, Target population reference LD estimates and Source-ancestry PRS weights. (B) **Three main steps** of the PTL-PRS framework, using the inputs in the orange boxes (numbered as in (A)) to fine-tune PRS weights of source population for the target population. **Step 0 (pseudosplitting)**: optional step to split the summary statistics data into training and validation sets. **Step 1 (training)**: gradient descent algorithm following TL-PRS procedures but with enhancements like blockwise early stopping (rightmost box in (B)) to optimize memory and time complexity. **Step 2 (evaluation)**: uses summary statistics to approximate the PRS-phenotype correlation (pseudo-*R*), guiding blockwise early stopping in Step 1 and hyperparameter tuning.

Step 1 is based on TL-PRS [9], while Steps 0 and 2 are adapted from the MegaPRS model [11].

**Step 0 (pseudosplitting)** is an optional step applied when only one GWAS summary statistics dataset is available. It splits the summary statistics data into training and validation sets. With these pseudosplit summary statistics along with target LD estimates and source PRS weights, **Step 1 (training)** is performed, following TL-PRS procedures but with enhancements such as blockwise early stopping (rightmost box in (B) of **Figure 1**) to optimize memory and time complexity. **Step 2 (evaluation)** involves the use of summary statistics to approximate the PRS-phenotype correlation (pseudo-*R*), guiding blockwise early stopping in Step 1 and hyperparameter tuning.

### STEP 0: Subsampling Pseudo-Summary Statistics

This step generates distinct summary statistics for training, validation, and testing (if necessary). It is achieved by emulating SNP marginal effect sizes (*γ*) through a random sampling technique [10]. The distribution used for this sampling, including variance estimation and detailed computational steps, is described in Supplementary Equation 3.

### STEP 1: Transfer learning of Polygenic Risk Scores

#### Recap of TL-PRS

TL-PRS [9] leverages transfer learning by initializing PRS weights from a source dataset and subsequently fine-tuning them via a gradient descent algorithm that is based on target ancestry data. The detailed equations are provided in Supplementary Equation 1.

#### Blockwise Early-stopping

We improved the early-stopping mechanism in TL-PRS by leveraging approximately independent linkage disequilibrium (LD) blocks [12]. While TL-PRS originally used LD blocks to parallelize training, we extended this approach to include parallelized model validation, further enhancing computational efficiency. Each LD block is treated as an independent unit for training and validation. After each iteration of coefficient update, we compute pseudo-R^2^ values for each block (see Step 2 and Supplementary Equation 2 for details). If the pseudo-R^2^ value for a given block decreases in subsequent iterations, coefficient updates for that block are terminated.

Unlike TL-PRS, which stores all intermediate coefficients across iterations and learning rates, PTL-PRS retains only the final set of coefficients per learning rate, significantly reducing storage usage. The final coefficients are concatenated across LD blocks, with each block potentially stopped at a different iteration because of blockwise early stopping. Additionally, pseudo-*R*^2^ values are computed blockwise and aggregated to select the optimal learning rate, eliminating redundant computations for LD blocks that have already converged.

### STEP 2: Calculation of Pseudo-*R*

To eliminate reliance on individual-level data for model evaluation, we adopted a pseudo-*R*^2^ implemented in MegaPRS [11]. This approach leverages standardized genotypic and phenotypic data to compute the Pearson correlation coefficient between observed and predicted phenotypes. The detailed formula for calculating pseudo-R^2^ is provided in Supplementary Equation 2.

### Computational Optimization

To increase computational efficiency, we reimplemented core functions in C++, including binary file reading and coefficient updating. PTL-PRS directly accesses PLINK (.bed) genotype files through an index-based search implemented in C++ using Rcpp [13], eliminating the need for intermediate txt file conversions required by TL-PRS. Using RcppParallel [14], we reimplemented the coefficient update function, which is the most time-consuming step, in C++ and parallelized it across LD blocks. To reduce memory usage, we divided the LD block input data, the main contributor to peak RAM consumption, into smaller chunks. These chunks are stored in queues, passed to the coefficient update function in parallel, and then removed from memory after processing.

## Results

We evaluated the performance of PTL-PRS by (1) comparing it with the original TL-PRS across eight phenotypes, (2) assessing the predictive accuracy for COVID-19 severity, and (3) measuring enhancements in time and memory efficiency. We tested four models—PTL-PRS-cs, PTL-PRS-lsum, TL-PRS-cs, and TL-PRS-lsum—categorized in Supplementary Table S1. The prefixes “PTL-PRS” and “TL-PRS” indicate whether pseudovalidation was applied, whereas the suffixes “-cs” and “-lsum” specify the underlying PRS method: PRS-cs [15] or Lassosum [16].

### Comparison of PTL-PRS and TL-PRS Performance

We evaluated the effects of PTL-PRS on eight phenotypes in South Asian ancestry: Type 2 Diabetes (T2D), High Density Lipoprotein (HDL), Low Density Lipoprotein (LDL), Body Mass Index (BMI), Height (HGT), Systolic Blood Pressure (SBP), Diastolic Blood Pressure (DBP), and Triglycerides (TG). These results were compared with those obtained with the TL-PRS method. Target ancestry summary statistics were derived from 6,588 South Asian UK Biobank participants, whereas source ancestry PRS weights were obtained from ExPRSweb [17] via the PGS Catalog [18], trained on UK Biobank White British and MGI European samples.

After reserving 10% of the data for testing, the remaining samples were pseudosplit into training and validation sets (9:1). Relative accuracy, defined as the percentage increase in *R*^2^ over the baseline European PRS model without fine-tuning, is illustrated in Supplementary Figure S1. The bar plots in dark blue and dark red represent the relative accuracy of PTL-PRS-cs and PTL-PRS-lsum, respectively, whereas the lighter-colored bars represent the corresponding TL-PRS results. The PTL-PRS and TL-PRS models exhibited comparable performance levels for each trait. Specifically, the PTL-PRS-cs and TL-PRS-cs models demonstrated average *R*^2^ improvements of 8.38% and 9.78%, respectively. Similarly, the PTL-PRS-lsum and TL-PRS-lsum models achieved average *R*^2^ improvements of 6.13% and 7.62%, respectively, across traits. Detailed performance metrics are provided in Supplementary Table S2.

### Improved Predictive Accuracy without Individual-level Data

We applied the PTL-PRS to construct polygenic risk scores (PRSs) for COVID-19 severity, comparing critically ill patients to those with mild symptoms in South Asian individuals using GenOMICC Release 2 GWAS summary statistics [19]. As individual-level validation or test data are unavailable, making TL-PRS inapplicable, we instead utilized publicly accessible summary statistics from South Asian populations (4,581 individuals; 788 cases) as the target data and those with European populations (48,880 individuals; 5,989 cases) to compute the source PRS. We performed two rounds of pseudosplitting to produce distinct training, validation, and test summary statistics at an 8:1:1 ratio. Since Lassosum requires individual-level or separate summary statistics for training and validation samples, we only used the PTL-PRS-cs model.

Owing to the lack of access to individual-level test data, the final performance was evaluated using pseudo-*R*^2^ . We note that its validity was assessed by comparing pseudo-*R*^2^ and true-*R*^2^ values across the eight previously mentioned traits, as shown in Supplementary Table S3. While pseudo-*R*^2^ values were slightly inflated, which is consistent with findings reported in PUMAS [10], the relative improvement patterns closely aligned with those of true-*R*^2^ , supporting pseudo-*R*^2^ as a reliable metric for evaluating relative performance improvements.

Supplementary Figure S2 presents the relative accuracy of the COVID-19 severity PRS, defined as the percentage ratio of the pseudo-*R*^2^ of the fine-tuned model to that of the baseline. To account for the stochastic nature of sample splitting, we repeated the procedure 25 times using different random seeds. These seeds induced variation in the distribution of marginal effect sizes during the pseudosplitting of the training and validation sets, helping to mitigate the variability from the pseudosplitting. Supplementary Figure S3 illustrates the running mean of the relative accuracy as the number of random seeds increases, with shaded regions indicating the standard error. As the number of seeds increases (in our case, beyond 15 random seeds), the running mean flattens, and the standard error decreases, suggesting that using multiple random seeds leads to more robust and reliable results. PTL-PRS-cs achieved an average increase of 6.22% in pseudo-*R*^2^ , with a standard deviation of 5.23. The detailed results are shown in Supplementary Table S4.

### Enhanced Computational Efficiency

We developed the R package *PTL*.*PRS*, which optimizes the original TL-PRS framework via blockwise early-stopping and computational optimization using C++. We benchmarked *PTL*.*PRS* against *TLPRS* (an R implementation of the original TL-PRS method) using 984,143 SNPs to predict LDL levels in 6,500 individuals of South Asian ancestry. Benchmarks were created on 3 CPU cores over 30 repeated runs. *PTL*.*PRS* achieved a median execution time of 25.6 minutes, corresponding to a 13.7-fold speedup over *TLPRS*, as shown in Supplementary Figure S4. Incorporating the additional pseudosplitting step for generating pseudo-summary statistics increased the median execution time by 8.01 minutes. In terms of resource efficiency, compared with *TLPRS, PTL*.*PRS* reduced total RAM usage by 8.15-fold, peak RAM usage by 1.48-fold, and the output file size by 7.15-fold.

## Conclusion

We developed PTL-PRS, an enhanced version of the TL-PRS framework that eliminates the need for individual-level data during model validation. This advancement broadens the framework’s applicability to settings where privacy concerns or data access limitations prevent the sharing of individual-level genotypes or phenotypes. Across eight common phenotypes, PTL-PRS demonstrated predictive accuracy comparable to that of TL-PRS while offering improved computational efficiency, positioning it as a robust and scalable alternative for data-limited contexts. When applied to COVID-19 severity data, the PTL-PRS yielded substantial gains in prediction accuracy. These results highlight the potential of PTL-PRS as a powerful tool for genetic risk prediction in scenarios with limited sample sizes and restricted data accessibility.

## Supporting information

Suuuuuplementary

## Data Availability

The summary statistics used in this paper are available in the public domain: UK Biobank (https://pheweb.org/UKB-TOPMed), PGS Catalog (https://www.pgscatalog.org) and GenOMICC (https://genomicc.org/data).

## Funding

This study was supported by the Brain Pool Plus (BP+) Program through the National Research Foundation of Korea (NRF) funded by the Ministry of Science and ICT [2020H1D3A2A03100666].

## Notes

### Competing Interest Statement

The authors have declared no competing interest.

### Author Declarations

Imputed genotype data from UKBiobank was used

## References

1. Choi, S.W., T.S.-H. Mak, and P.F. O’Reilly, Tutorial: a guide to performing polygenic risk score analyses. Nature Protocols, 2020. 15(9): p. 2759–2772.

2. Henches, L., et al., Polygenic risk score prediction accuracy convergence. 2023, Genetics.

3. Torkamani, A., N.E. Wineinger, and E.J. Topol, The personal and clinical utility of polygenic risk scores. Nature Reviews Genetics, 2018. 19(9): p. 581–590.

4. Khera, A.V., et al., Genome-wide polygenic scores for common diseases identify individuals with risk equivalent to monogenic mutations. Nature Genetics, 2018. 50(9): p. 1219–1224.

5. Lambert, S.A., G. Abraham, and M. Inouye, Towards clinical utility of polygenic risk scores. Human Molecular Genetics, 2019. 28(R2): p. R133–R142.

6. Martin, A.R., et al., Clinical use of current polygenic risk scores may exacerbate health disparities. Nature Genetics, 2019. 51(4): p. 584–591.

7. Weissbrod, O., et al., Leveraging fine-mapping and multipopulation training data to improve cross-population polygenic risk scores. Nature Genetics, 2022. 54(4): p. 450–458.

8. Lewis, C.M. and E. Vassos, Polygenic risk scores: from research tools to clinical instruments. Genome Medicine, 2020. 12(1): p. 44.

9. Zhao, Z., et al., The construction of cross-population polygenic risk scores using transfer learning. The American Journal of Human Genetics, 2022. 109(11): p. 1998–2008.

10. Zhao, Z., et al., PUMAS: fine-tuning polygenic risk scores with GWAS summary statistics. Genome Biology, 2021. 22(1): p. 257.

11. Zhang, Q., et al., Improved genetic prediction of complex traits from individual-level data or summary statistics. Nature Communications, 2021. 12(1): p. 4192.

12. Berisa, T. and J.K. Pickrell, Approximately independent linkage disequilibrium blocks in human populations. Bioinformatics, 2016. 32(2): p. 283–285.

13. Eddelbuettel, D. and R. François, Rcpp : Seamless R and C++ Integration. Journal of Statistical Software, 2011. 40(8).

14. Jj, A., et al., RcppParallel: Parallel Programming Tools for ‘Rcpp’. 2025.

15. Ge, T., et al., Polygenic prediction via Bayesian regression and continuous shrinkage priors. Nature Communications, 2019. 10(1): p. 1776.

16. Mak, T.S.H., et al., Polygenic scores via penalized regression on summary statistics. Genetic Epidemiology, 2017. 41(6): p. 469–480.

17. Ma, Y., et al., ExPRSweb: An online repository with polygenic risk scores for common health-related exposures. The American Journal of Human Genetics, 2022. 109(10): p. 1742–1760.

18. Lambert, S.A., et al., The Polygenic Score Catalog as an open database for reproducibility and systematic evaluation. Nature Genetics, 2021. 53(4): p. 420–425.

19. Kousathanas, A., et al., Whole-genome sequencing reveals host factors underlying critical COVID-19. Nature, 2022. 607(7917): p. 97–103.

