## Supplementary material for "PTL-PRS: an R package for transfer learning of polygenic risk scores with pseudovalidation": Suuuuuplementary

Supplementary Data

Bokeum Cho^1^, Seunggeun Lee^1,*^

^1^ Graduate School of Data Science, Seoul National University, Seoul, South Korea

**Table of Content**

Supplementary Figure S1. Relative accuracy of PTL-PRS and TL-PRS for different traits.

- Supplementary Figure S2. Relative accuracy of PTL-PRS-cs for COVID-19 severity across different random seeds, using pseudo-*R*.
- Supplementary Figure S3. Running mean of relative accuracy as the number of random seeds increases.
- Supplementary Figure S4. Comparison of computational cost for training PTL-PRS and TL-PRS.
- Supplementary Equation 1. Recap of TL-PRS
- Supplementary Equation 2. Formula for pseudo-*R*
- Supplementary Equation 3. Formula for pseudosplitting
- Supplementary Table S1. 4 Types of models used in analysis
- Supplementary Table S2. Baseline $R^{2}$, model $R^{2}$, and relative accuracy for different methods and traits
- Supplementary Table S3. Baseline $R^{2}$, model $R^{2}$, and relative accuracy for different methods and traits, using both true and pseudo-$R^{2}$ metrics
- Supplementary Table S4. Baseline pseudo-$R^{2}$, model pseudo-$R^{2}$, and relative accuracy of PTL-PRS-cs across different seeds used for pseudosplitting training and validation sets

Supplementary Figure S1. Relative accuracy of PTL-PRS and TL-PRS for different target phenotypes. The bar plots in dark blue and dark red represent the relative accuracy of PTL-PRS-cs and PTL-PRS-lsum, respectively, while the lighter-colored bars show the corresponding TL-PRS results.

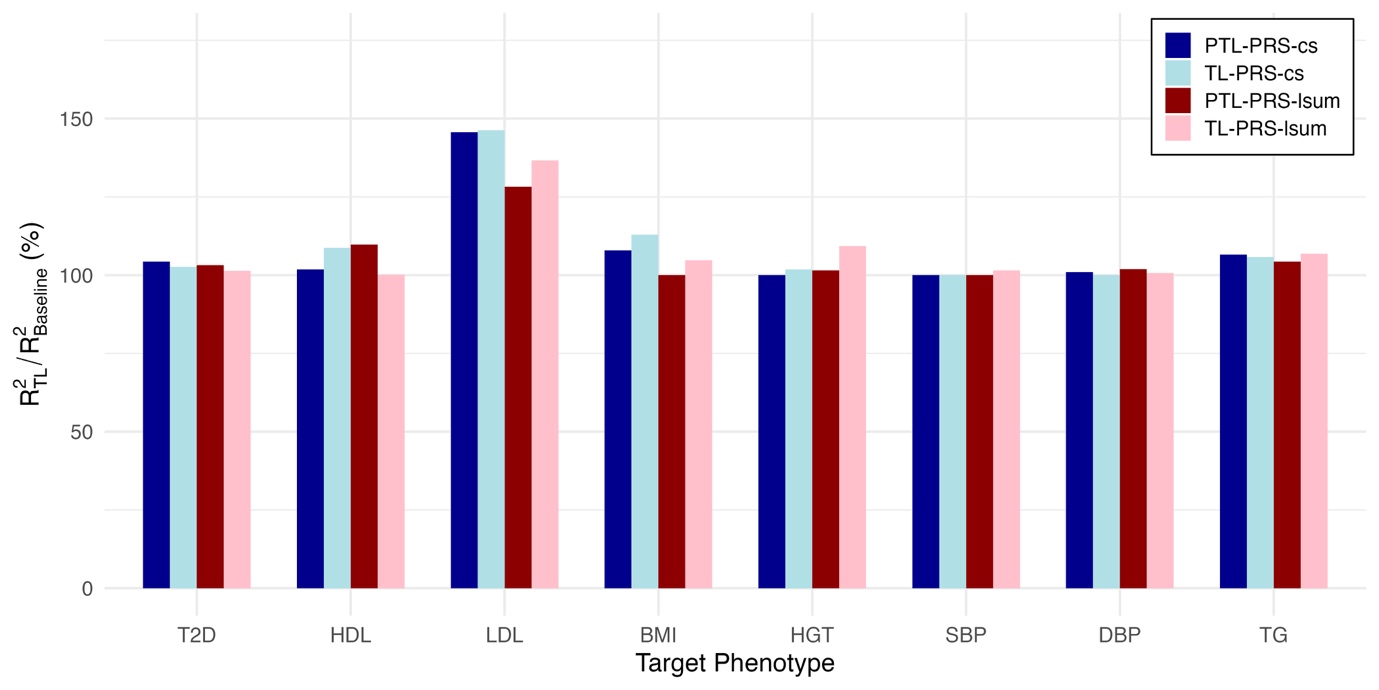

Supplementary Figure S2. Relative accuracy of PTL-PRS-cs for COVID-19 severity across different random seeds, using pseudo-R. Each dot represents the relative accuracy for a different random seed.
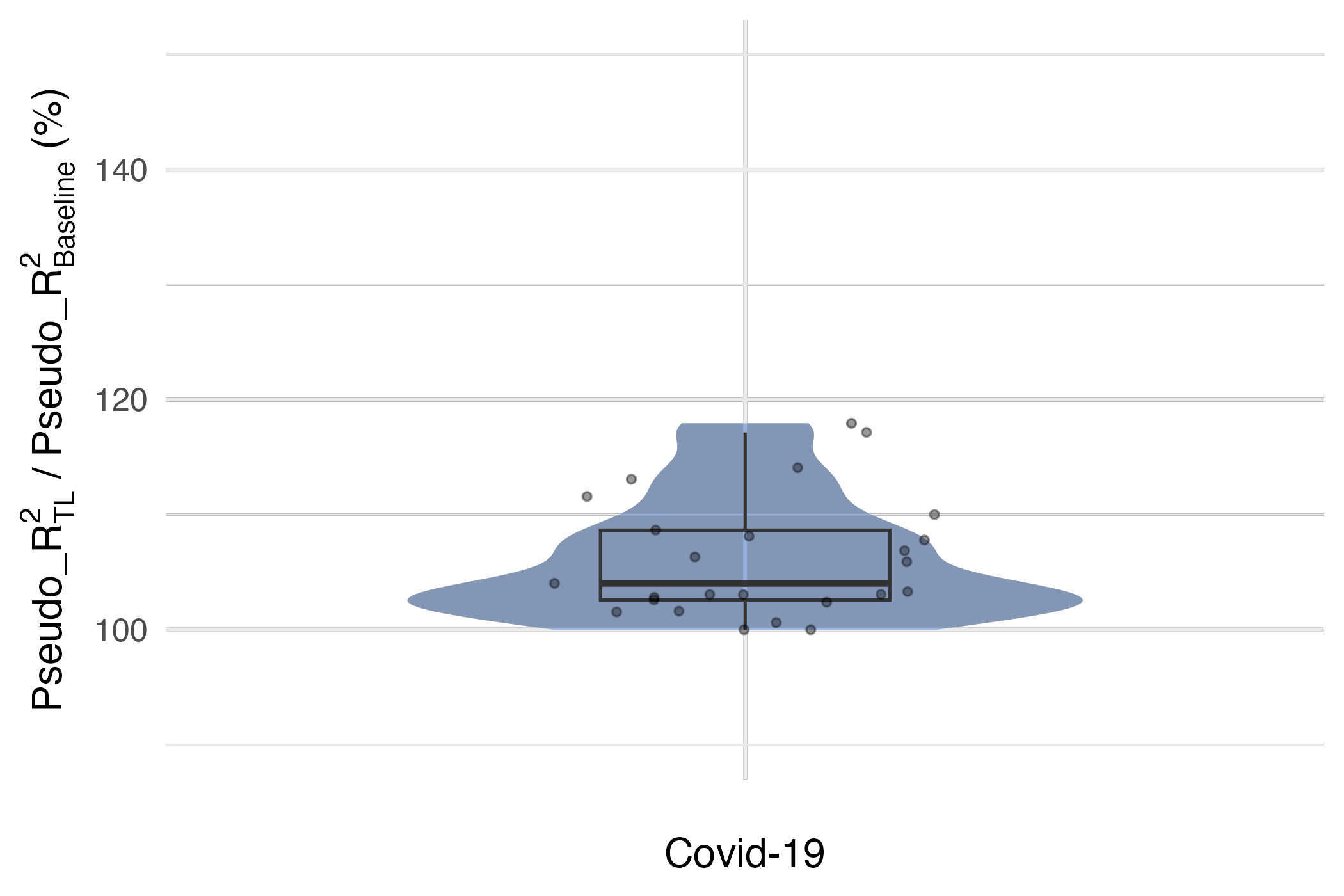

Supplementary Figure S3. Running mean of relative accuracy as the number of random seeds increases. The shaded area (ribbon) represents ±1 standard error around the mean.

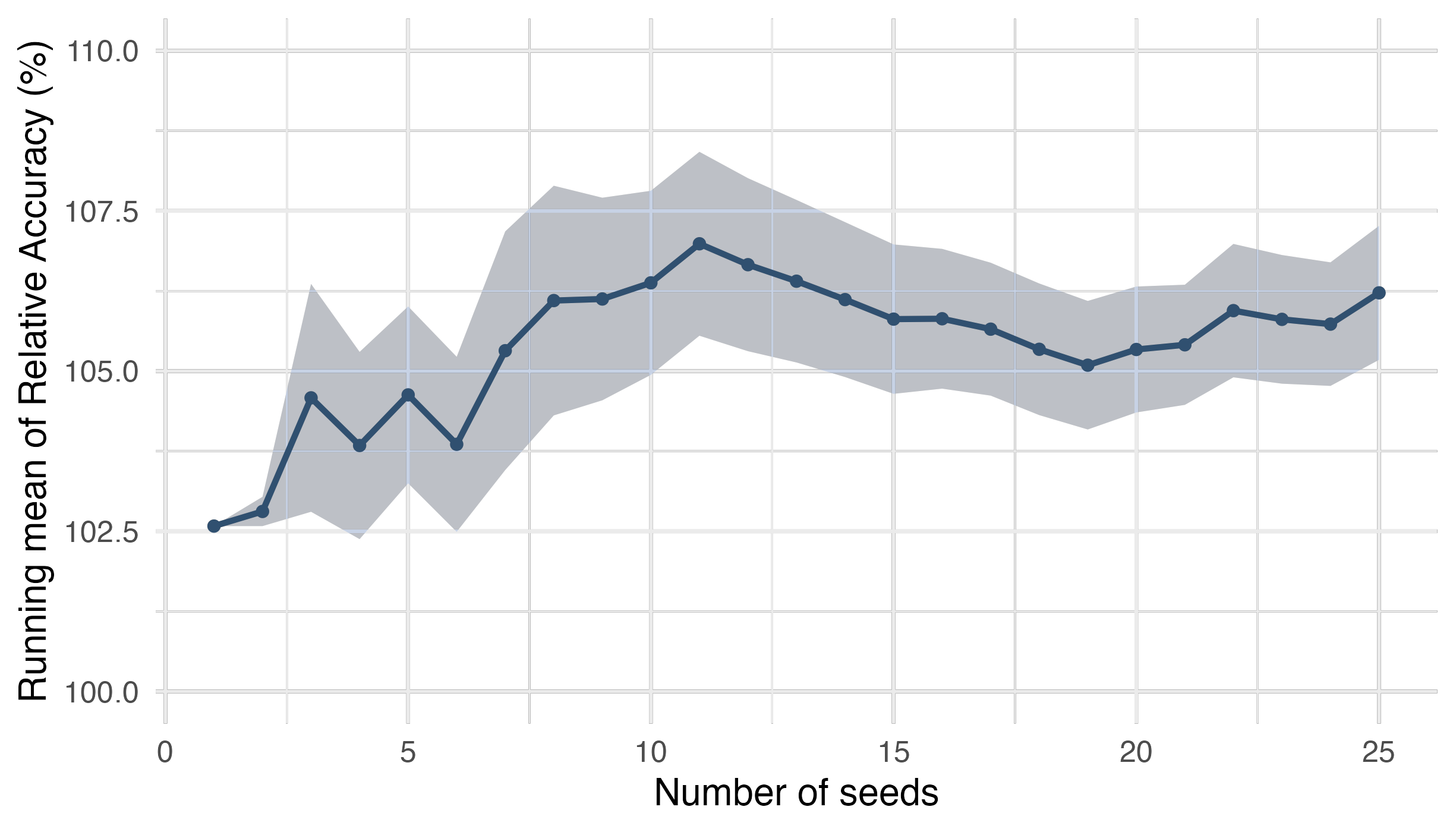

Supplementary Figure S4. Comparison of computational cost for training PTL-PRS and TL-PRS: execution time in hours (top left), total output size in MB (top right), total RAM usage in MB (bottom left) and peak RAM usage in GB (bottom right). Benchmarking was conducted using three cores of an AMD EPYC 7542 32-core processor.

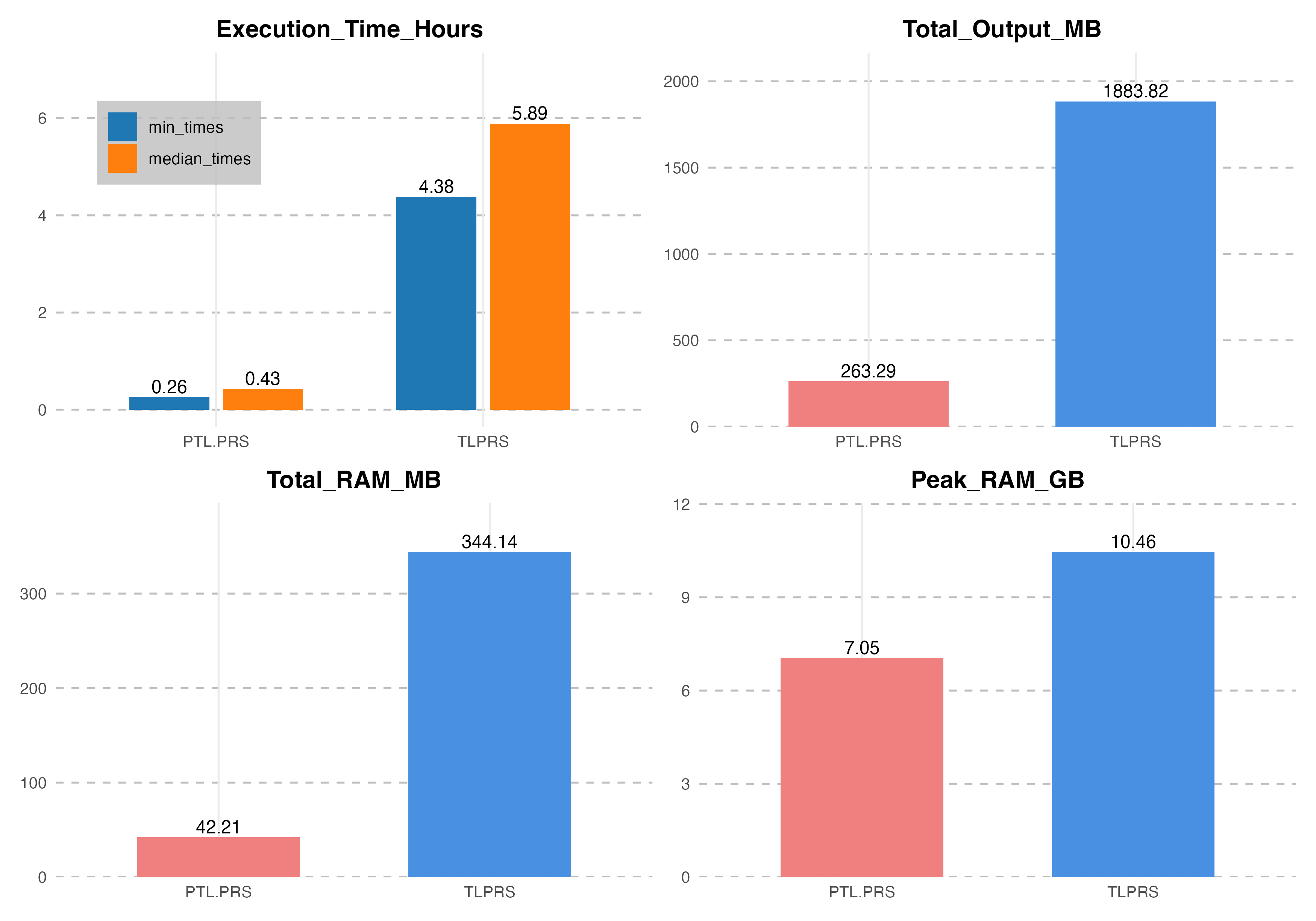

**Supplementary Equation 1. Recap of TL-PRS**

A Polygenic Risk Score (PRS) is formulated as the aggregate of estimated effects across all genetic variants relevant to a specific phenotype. For a given individual denoted as *i*, the PRS can be calculated as following:

$$PRS_{i}=\sum_{j=1}^{M} \hat{\beta_{j}}G_{ij}$$

where M represents the total number of variants, $G_{ij}$ denotes the genotype of genetic variant *j*, and $\hat{\beta_{j}}$ is the marginal effect size of the variant *j*.

$$\begin{aligned} \boldsymbol{Y}=\sum_{j=1}^{M} G_{j}\beta_{j}+\varepsilon=\sum_{j=1}^{M} G_{j}\left( \beta_{j}^{pre}+\tau_{j} \right)+\varepsilon\boldsymbol{\#}\left( 1 \right) \end{aligned}$$

$$\begin{aligned} Loss=\left( \boldsymbol{Y}-\sum_{j=1}^{M} G_{j}\beta_{j} \right)^{2}\#\left( 2 \right) \end{aligned}$$

$$\begin{aligned} \beta_{j}^{\left( r+1 \right)}=\beta_{j}^{\left( r \right)}-\alpha^{'}\frac{\partial Loss}{\partial\beta_{j}}=\beta_{j}^{\left( r \right)}+2\alpha^{'}G_{j}^{T}\left( \boldsymbol{Y}-\boldsymbol{G}\beta^{\left( r \right)} \right)\#\left( 3 \right) \end{aligned}$$

The equation 1 refers to the model form, equation 2 shows the loss function, and the equation 3 is a formula to estimate the next coefficient of variant *j*, given the current estimate $\beta_{j}^{\left( r \right)}$. During the model fitting phase, summary statistics of the target group can be utilized to estimate $\boldsymbol{G}^{T}\boldsymbol{Y}$ and$\boldsymbol{G}^{T}\boldsymbol{G}$ can be derived from the public reference datasets, such as the 1000 Genomes Project. The learning rate $\alpha=2\alpha^{'}$ can be chosen as parameter among the values the user offers, based on the validation dataset to optimize prediction accuracy. The default grid of learning rate is $min\left( \frac{1,10,100,1000}{n\left( SNPs \right)},1 \right)$.

**Supplementary Equation 2. Formula for pseudo-*R***

Assuming that both the genotype and phenotype data are standardized, the correlation between observed and predicted phenotypes for the individuals used in computing the validation or test summary statistics can be expressed as:

$$R=\frac{\beta^{'T}\boldsymbol{X}^{T}\boldsymbol{Y}}{\sqrt{n\left( \beta^{'T}\boldsymbol{X}^{T}\boldsymbol{X}\beta^{'} \right)}} ,$$

where $\beta^{'}$ represents the vector of estimated effect sizes for a training model and $\boldsymbol{X}$ is the standardized form of genotype matrix $\boldsymbol{G}$.

Analogous to the model-fitting phase in TL-PRS, $\boldsymbol{X}^{T}\boldsymbol{Y}$ can be estimated with SNP-phenotype correlations ($\hat{\rho}$) from the validation summary statistics of the target phenotype, and $\boldsymbol{X}^{T}\boldsymbol{X}$ can be substituted with SNP-wise correlation ($\hat{r}$) from public reference dataset.

**Supplementary Equation 3. Formula for pseudosplitting**

The goal is to mimic marginal SNP effect sizes $\boldsymbol{\gamma}=\left( \gamma_{1},\gamma_{2},\ldots,\gamma_{m} \right)^{T}=G^{T}Y/n$, which equals to the correlation between SNP *j* and the phenotype, of distinct sample sets for train and test. Here, we assume $G$ and $Y$ are centered. $V$ refers to the variance of $G^{T}Y$. We obtain the following estimates from training (A) and test (B) sets:

- Estimate of $\boldsymbol{\gamma}$ from $n_{A}$ training samples:

$$\frac{\boldsymbol{G}_{\boldsymbol{A}}^{\boldsymbol{T}}\boldsymbol{Y}_{\boldsymbol{A}}}{n_{A}}\sim\mathcal{N}\left( \frac{\boldsymbol{G}^{\boldsymbol{T}}\boldsymbol{Y}}{n}, \frac{n_{B}}{n_{A}}\cdot\frac{\boldsymbol{V}}{n} \right)$$

- Estimate of $\boldsymbol{\gamma}$ from $n_{B}$ test samples:

$$\frac{G_{B}^{T}Y_{B}}{n_{B}}=G^{T}Y-\frac{G_{A}^{T}Y_{A}}{n_{B}}$$

To account for linkage disequilibrium, we follow Zhu and Stephens [1] and set:

$$V=G^{T}G$$

If $X^{'}$ denotes the standardized genotypes of a reference panel of size $n^{'}\times m$, we approximate $V$ by:

$$V \approx X^{'T}X^{'} \frac{n}{n^{'}}$$

Hence, we achieve the desired sampling by defining:

$$\frac{G_{A}^{T} Y_{A}}{n_{A}}=\frac{G^{T}Y}{n} + \sqrt{\frac{n_{B}}{n_{A}}} \frac{X^{'T}}{\sqrt{n^{'}}} g ,$$

where $g$ is a vector of length $n'$ with elements drawn from a standard Gaussian distribution.

Supplementary Table S1. 4 Types of models used in analysis

|  |  | Pseudovalidation | |
| --- | --- | --- | --- |
|  |  | X | O |
| Baseline PRS | PRS-cs | TL-PRS-cs | PTL-PRS-cs |
|  | Lassosum | TL-PRS-Lsum | PTL-PRS-Lsum |

Supplementary Table S2. Baseline $\boldsymbol{R}^{\boldsymbol{2}}$, model $\boldsymbol{R}^{\boldsymbol{2}}$, and relative accuracy for different methods and traits

|  | PTL-PRS-cs | | | TL-PRS-cs | | |
| --- | --- | --- | --- | --- | --- | --- |
| Pheno | Baseline $R^{2}$ | Model $R^{2}$ | Rel. Acc.(%) | Baseline $R^{2}$ | Model $R^{2}$ | Rel. Acc.(%) |
| T2D | 0.01589 | 0.01658 | 104.35 | 0.01069 | 0.01097 | 102.60 |
| HDL | 0.09793 | 0.09967 | 101.77 | 0.02256 | 0.02454 | 108.78 |
| LDL | 0.02452 | 0.03571 | 145.66 | 0.01267 | 0.01853 | 146.28 |
| BMI | 0.03658 | 0.03945 | 107.83 | 0.00743 | 0.00839 | 113.01 |
| HGT | 0.07327 | 0.07327 | 100.00 | 0.03864 | 0.03935 | 101.85 |
| SBP | 0.05087 | 0.05087 | 100.00 | 0.02258 | 0.02258 | 100.00 |
| DBP | 0.05018 | 0.05065 | 100.94 | 0.01402 | 0.01402 | 100.00 |
| TG | 0.04501 | 0.04793 | 106.49 | 0.01134 | 0.01199 | 105.80 |
|  | PTL-PRS-lsum | | | TL-PRS-lsum | | |
| Pheno | Baseline $R^{2}$ | Model $R^{2}$ | Rel. Acc.(%) | Baseline $R^{2}$ | Model $R^{2}$ | Rel. Acc.(%) |
| T2D | 0.02365 | 0.02440 | 103.18 | 0.02455 | 0.02488 | 101.36 |
| HDL | 0.03368 | 0.03696 | 109.74 | 0.06819 | 0.06825 | 100.09 |
| LDL | 0.01779 | 0.02281 | 128.24 | 0.01878 | 0.02565 | 136.58 |
| BMI | 0.03337 | 0.03338 | 100.03 | 0.03481 | 0.03645 | 104.71 |
| HGT | 0.03135 | 0.03182 | 101.49 | 0.03926 | 0.04288 | 109.21 |
| SBP | 0.04668 | 0.04671 | 100.05 | 0.05012 | 0.05089 | 101.52 |
| DBP | 0.03454 | 0.03520 | 101.92 | 0.02147 | 0.02161 | 100.66 |
| TG | 0.03122 | 0.03258 | 104.38 | 0.03099 | 0.03310 | 106.82 |

Supplementary Table S3. Baseline $\boldsymbol{R}^{\boldsymbol{2}}$, model $\boldsymbol{R}^{\boldsymbol{2}}$, and relative accuracy for different methods and traits, using both true and pseudo-$\boldsymbol{R}^{\boldsymbol{2}}$ metrics

| Method | Pheno | Baseline $R^{2}$ | Model $R^{2}$ | Rel. Acc.(%) | Baseline pseudo-$R^{2}$ | Model  pseudo-$R^{2}$ | Rel. Acc.(%) |
| --- | --- | --- | --- | --- | --- | --- | --- |
| PTL-PRS-cs | T2D | 0.01589 | 0.01658 | 104.35 | 0.01764 | 0.01778 | 100.76 |
|  | HDL | 0.09793 | 0.09967 | 101.77 | 0.09415 | 0.09491 | 100.81 |
|  | LDL | 0.02452 | 0.03571 | 145.66 | 0.02333 | 0.02487 | 106.58 |
|  | BMI | 0.03658 | 0.03945 | 107.83 | 0.05562 | 0.05612 | 100.90 |
|  | HGT | 0.07327 | 0.07327 | 100.00 | 0.19596 | 0.19852 | 101.31 |
|  | SBP | 0.05087 | 0.05087 | 100.00 | 0.09254 | 0.09254 | 100.00 |
|  | DBP | 0.05018 | 0.05065 | 100.94 | 0.06685 | 0.06685 | 100.00 |
|  | TG | 0.04501 | 0.04793 | 106.49 | 0.08250 | 0.08586 | 104.05 |
| PTL-PRS-lsum | T2D | 0.02365 | 0.02440 | 103.18 | 0.03025 | 0.03087 | 102.04 |
|  | HDL | 0.03368 | 0.03696 | 109.74 | 0.07196 | 0.07537 | 104.74 |
|  | LDL | 0.01779 | 0.02281 | 128.24 | 0.03070 | 0.03490 | 113.68 |
|  | BMI | 0.03337 | 0.03338 | 100.03 | 0.05108 | 0.05122 | 100.28 |
|  | HGT | 0.03135 | 0.03182 | 101.49 | 0.17854 | 0.17927 | 100.41 |
|  | SBP | 0.04668 | 0.04671 | 100.05 | 0.09292 | 0.09325 | 100.36 |
|  | DBP | 0.03454 | 0.03520 | 101.92 | 0.04642 | 0.04674 | 100.67 |
|  | TG | 0.03122 | 0.03258 | 104.38 | 0.07743 | 0.07805 | 100.79 |

Supplementary Table S4. Baseline pseudo-$\boldsymbol{R}^{\boldsymbol{2}}$, model pseudo-$\boldsymbol{R}^{\boldsymbol{2}}$, and relative accuracy of PTL-PRS-cs across different seeds used for pseudosplitting training and validation sets

| Seed | Baseline pseudo-$R^{2}$ | Model pseudo-$R^{2}$ | Relative Accuracy (%) |
| --- | --- | --- | --- |
| 10 | 0.0013887 | 0.0014245 | 102.58 |
| 15 |  | 0.0014309 | 103.04 |
| 20 |  | 0.0015016 | 108.13 |
| 40 |  | 0.0014110 | 101.61 |
| 55 |  | 0.0014969 | 107.79 |
| 60 |  | 0.0013887 | 100.00 |
| 65 |  | 0.0015843 | 114.08 |
| 75 |  | 0.0015495 | 111.58 |
| 90 |  | 0.0014765 | 106.32 |
| 95 |  | 0.0015088 | 108.65 |
| 125 |  | 0.0015704 | 113.08 |
| 130 |  | 0.0014313 | 103.07 |
| 140 |  | 0.0014347 | 103.31 |
| 145 |  | 0.0014218 | 102.39 |
| 150 |  | 0.0014101 | 101.54 |
| 160 |  | 0.0014707 | 105.91 |
| 165 |  | 0.0014311 | 103.06 |
| 170 |  | 0.0013884 | 100.00 |
| 175 |  | 0.0013973 | 100.62 |
| 195 |  | 0.0015276 | 110.00 |
| 200 |  | 0.0014841 | 106.87 |
| 250 |  | 0.0016268 | 117.15 |
| 280 |  | 0.0014277 | 102.81 |
| 290 |  | 0.0014445 | 104.02 |
| 300 |  | 0.0016379 | 117.94 |
| **AVG** |  | **0.0014751** | **106.22** |

**References**

1.	Zhu, X. and M. Stephens, *Bayesian large-scale multiple regression with summary statistics from genome-wide association studies.* The Annals of Applied Statistics, 2017. **11**(3).
